# Design and validation of an automated radiation therapy treatment planning approach for locally advanced lung cancer

**DOI:** 10.1101/2022.09.30.22280584

**Authors:** Joel A. Pogue, Carlos E. Cardenas, Joseph Harms, Michael H. Soike, Adam J. Kole, Craig S. Schneider, Christopher Veale, Richard Popple, Jean-Guy Belliveau, Andrew M. McDonald, Dennis N. Stanley

**Affiliations:** Department of Radiation Oncology, University of Alabama at Birmingham; University of Alabama at Birmingham Institute for Cancer Outcomes and Survivorship

## Abstract

**Purpose:** Radiation therapy planning for locally-advanced non-small cell lung cancer (NSCLC) is challenging due to the balancing of target coverage and organs-at-risk (OAR) sparing. Using the Varian Ethos Treatment Planning System (TPS), we developed a methodology to automatically generate efficient, high-quality treatment plans for locally-advanced lung cancer patients.

**Methods and Materials:** Fifty patients previously treated with Eclipse-generated plans for inoperable Stage IIIA-IIIC NSCLC were included in this Institutional Review Board (IRB)-approved retrospective study. Fifteen patients were used to iteratively optimize an Ethos TPS planning template, and the remaining thirty-five patients had plans automatically generated without manual intervention using the created template. Ethos and Eclipse plan quality was then assessed using 1) standard dose volume histogram (DVH) metrics, 2) adherence to clinical trial objectives, and 3) radiation oncologist qualitative review.

**Results:** Ethos-generated plans showed improved primary and nodal planning target volume (PTVp and PTVn, respectively) V100% and V95% coverage (p<0.001) and reduced PTVp Dmax values (p=0.023). Furthermore, the Ethos template-generated plans had lower spinal cord Dmax, lungs V5Gy, and heart V25Gy, V30Gy, and V45Gy values (p≤0.021). However, Ethos esophagus metrics (mean, V35Gy, V50Gy, Dmax) and brachial plexus metrics (Dmax) were greater than Eclipse (p≤0.008), but were still clinically acceptable. A large majority (80%) of automatically generated plans had entirely “per protocol” or “variation acceptable” metrics. Three radiation oncologists qualitatively scored the Ethos plans; 78% of plans were scored as clinically acceptable during physician evaluation, with zero plans receiving scores requiring major changes.

**Conclusions:** A standard Ethos template generated lung cancer radiotherapy plans with greater target coverage, increased spinal cord, heart, and lung V5Gy sparing, but increased esophagus and brachial plexus dose, compared to manually generated Eclipse plans. This template elucidates an efficient approach for generating automated, high quality lung radiation therapy treatment plans.

## Introduction

While significant strides have been made towards automation in radiation oncology, radiation therapy (RT) treatment planning currently remains a largely manual process in which dosimetrists, physicists, and physicians iteratively develop plans on a patient-by-patient basis. This method of iterative planning costs significant departmental resources such as time required to train personnel and develop quality treatment plans.^1^ Manually generated plan quality is highly heterogeneous due to individual planner skill and experience. ^2-4^ Several factors affect the quality of radiotherapy plans including planner and Radiation Oncologist’s choice of optimization dose and priority values, planning time limitations, and the extent optimization structures and normal tissue constraints are utilized to limit dose outside the tumor.

For these reasons, automated planning approaches have been investigated. Automated planning aims to decrease inter-plan variation and planning time while maintaining or improving plan quality. ^5^ This is typically achieved through standardization of structures of interest, optimization techniques, and priority values. Specific approaches include, but are not limited to, knowledge-based planning (KBP), ^6-8^ multi-criteria optimization, ^9-11^ and template based planning.^12, 13^ To this end, the Ethos Adaptive RT platform (Varian Medical Systems, Palo Alto, CA) has been designed with an automated treatment planning system (TPS) that generates plans from user-provided templates.^14^

While the Ethos platform is relatively new, several aspects of the Ethos system have already been investigated. Mao et al. have shown that adaptive therapy on the Ethos platform significantly improves target coverage and reduces normal tissue dose.^15^ Other studies have demonstrated that Ethos daily adaptive prostate and abdominal SBRT plans generally result in higher target coverage and a reduced risk of exceeding clinical OAR thresholds compared to scheduled plans, and that the Ethos intensity modulated radiation therapy (IMRT) optimizer generates similar or better quality pelvis RT plans compared to manually generated Eclipse volumetric modulated arc therapy (VMAT) techniques.^16-19^ However, there has been limited investigation of Ethos thoracic RT plan quality. As the Ethos system introduces a new TPS, and the TPS drives RT plan generation, a quantitative understanding of the differences in dose distributions between conventional linear accelerator-based treatments and Ethos TPS treatments is needed. The primary endpoint of this retrospective treatment planning study is to develop an automated treatment planning approach for creating high quality plans in the Ethos workspace. As a secondary endpoint, Ethos-generated plans will be quantitatively compared to plans generated by skilled dosimetrists in Eclipse and treated on a conventional linear accelerator, then qualitatively evaluated by experienced radiation oncologists.

## Methods and Materials

### Patient and volume description

Fifty patients previously treated at our institution for inoperable stage IIIA-IIIC non-small cell lung cancer (NSCLC) between 2019 and 2022 were randomly selected for this Institutional Review Board (IRB-1207033005) approved study. Patients were required to meet inclusion criteria for the prospective ARTIA-Lung clinical trial (Daily Adaptive vs Non-Adaptive External Beam Radiation Therapy with Concurrent Chemotherapy for Locally Advanced Non-Small Cell Lung Cancer: A Prospective Randomized Trial of an Individualized Approach for Toxicity Reduction; clinicaltrials.gov identifier NCT05488626). Accordingly, patients were excluded if they had contralateral hilar or supraclavicular lymph nodes involvement, or distant metastases. All patients were immobilized according to institutional protocol and simulated head-first supine using a Phillips Brilliance big bore CT scanner, if patient mobility allowed. Simulation CT scans extended from the inferior aspect of the cricoid through the entire liver using a slice thickness of 2-3mm. As per thoracic planning CT protocol, the entire body surface was inside the field of view, and all patients received respiratory-correlated 4D-CT.

Targets and normal structures were generally delineated by the treating physician consistent with the Radiation Therapy Oncology Group (RTOG) 1106 study.^20^ Positron emission tomography CT (PET-CT) images were registered with the averaged 4D-CT to aid in delineation of primary and nodal gross tumor volumes (GTVp and GTVn, respectively), when available. Patients were treated free-breathing or with phase-based gating based on 4D motion assessment. Internal gross tumor volumes (iGTV) were defined as the union of GTVs drawn individually on each phase.

Primary and nodal clinical target volumes (CTVp and CTVn, respectively) were typically generated by adding isotropic 5mm or 7mm margins to the iGTVs, with manual cropping of the CTV at natural barriers to tumor invasion. As an example, should an isotropic expansion in the level 8 paraesophageal region infringe into the vertebral body, it was modified to remove infringement into the barrier. Primary and nodal PTVs (PTVp and PTVn, respectively) were generated by adding margins between 5mm and 10mm to the CTVs, most commonly 5mm isotropically (n=38). Total planning target volumes (PTV) ranged from 95.1cm^3^ to 1245.6cm^3^, with a mean value of 504.5cm^3^. All patients were prescribed 60Gy in thirty fractions. When plans were initially calculated in Eclipse with the anisotropic analytical algorithm (AAA, version 13.6.23, Varian Medical Systems), the prescription was normalized so that 95% of the PTV receives prescription dose, except in cases where organ at risk (OAR) dose was near tolerance. On average, 94.2% of the PTV volume received 60Gy.

### Treatment planning

Patients were treated with IMRT using 6-10 fields (n=6) or VMAT using 2-3 arcs (n=44) in the clinical, manually generated AAA treatment plans. To the contrary, the Ethos TPS calculates dose using AcurosXB (AXB, Varian Medical Systems) which is a Grid-Based Boltzmann Solver (GBBS) algorithm. Due to systematic differences between AXB and AAA when treating lung cancer, all previously treated clinical Eclipse plans were recalculated using AXB (version 15.5.11) with heterogeneity correction on and dose to medium reporting mode selected.^21-23^ Recalculations preserved the monitor units, beam geometries, and field weightings used in the clinically delivered plans. A 2mm calculation grid and 1.8° control point spacing were utilized. Patients were treated using 6MV flattened beams on a Varian Clinac 21IX and TrueBeam STX, each having a maximum dose rate of 600 MU/min and equipped with millennium 120 MLCs.^24^

The Ethos TPS (version 1.1, Varian Medical Systems) is designed for CBCT-guided adaptive radiation therapy delivered on a Halcyon rotational linear accelerator. The Halcyon utilizes a 6MV flattening filter free (FFF) beam which has a maximum dose rate of 800 MU/min and employs double stacked MLC banks as its primary form of collimation.^25^ For Ethos plan generation, all Eclipse structure sets were anonymized and imported into the Ethos Treatment Management system (version 02.01.00, Varian Medical Systems). Plans were calculated using AXB (version 16.1.0) with heterogeneity correction on and dose to medium reporting mode selected. A 2.5mm calculation grid and 2.0° control point spacing were utilized.

When replanning in Ethos, patients were randomly assigned to either the tuning (n=15) or validation (n=35) cohort until the tuning cohort had evenly distributed tumor laterality (left/right), ensuring the resulting template was robust to tumor location. The remaining thirty-five patient validation cohort was deemed of sufficient size to test the difference between Ethos and Eclipse plans with high confidence. Plans in the tuning cohort were used to iteratively develop and fine-tune an automated Ethos planning template. The final template was then used to generate plans for all fifty patients. Plans created using the template were evaluated “as-is” after the initial optimization (i.e. no further planning or normalization). Optimization priorities and dosimetric objectives were established based on the ARTIA-Lung clinical trial, shown in Supplemental Materials Table A1.

The Ethos TPS utilizes the intelligent optimization engine (IOE) to automatically generate multiple plans from each treatment intent using pre-defined beam geometries which are each optimized according to the same clinical priority template.^26^ Ethos template planning objectives increase in priority with ascending order in the dose preview workspace, rather than by a cost function that varies with assigned priority number, as in Eclipse. For consistency with the ARTIA-Lung clinical trial protocol, Ethos priority 1 (most important) and priority 2 (very important) objectives were utilized in this study. The pre-defined geometries selected for this work include equidistant 9-field and 12-field IMRT plans, an ipsilateral 7-field IMRT plan, 2 and 3 full-arc VMAT plans, and a 2 half-arc (180-degree arc span) VMAT plan. In this study, the optimal plan was selected for each patient according to the ARTIA-Lung metrics and hierarchy.

For final dose reporting, Ethos plans were exported and analyzed in Eclipse. Average and standard deviation values for all ARTIA-Lung trial metrics, and all pertinent additional Quantitative Analysis of Normal Tissue Effects (QUANTEC) metrics,^27^ were generated based on the recommendations of Amdur et al.^28^ Eclipse AAA, Eclipse AXB, and Ethos plan metrics and dose volume histograms (DVH) were extracted using the Eclipse Scripting Application Programming Interface. The Wilcoxon paired, non-parametric test was utilized to test the difference between Eclipse and Ethos generated plan metrics for all objectives.^29^ P-values were calculated, without removal of outliers, using the SciPy library in Python.

### Qualitative evaluation

Three board certified radiation oncologists with extensive experience treating lung cancer participated in plan reviews. All physicians initially reviewed five plans together to standardize the review process and normalize plan scoring. Each physician then independently reviewed thirty of the remaining forty-five plans, ensuring each of the fifty plans was reviewed by at least two physicians. The physicians were not provided additional clinical information regarding the cases and purely reviewed plan quality with anonymous patient identifiers. The physicians were not informed by physicists of the ARTIA-Lung clinical trial constraints, were not shown the template utilized for plan generation, and were not informed that plans had been calculated with AXB as opposed to AAA, which is the clinical standard. Instead, they judged plan quality according to their unique interpretation of the scoring criteria outlined below,

**5 - Use as-is**. Clinically acceptable plan that could be used for treatment without change.

**4 - Minor edits that are unnecessary**. Reviewer would recommend changes but considers current plan acceptable for treatment.

**3 - Minor edits that are necessary**. Reviewer would require changes prior to treatment and the changes, in the judgment of the reviewer, can be implemented by minimal editing of the objectives.

**2 - Major edits**. Reviewer would require changes prior to treatment and the changes in the judgment of the reviewer would require significant modification of the objectives.

**1 - Unusable**. The plan quality is so poor that it is deemed unsafe to deliver, i.e. would likely result in harm to the patient.

## Results

### Planning template

A total of 129 initial intents and intent revisions were created throughout this analysis (94 for the tuning cohort, 35 for the validation cohort), resulting in the calculation and subsequent evaluation of 774 unique radiation plans. This volume of intents was necessary because many of the ARTIA-Lung planning objectives were not met for the fifteen-patient tuning cohort when plans were directly optimized using the ARTIA-Lung objectives in the absence of optimization structures. To guide optimization, avoidance structures, as defined in the optimization template found in Table 1, were created for any normal tissues that are potentially proximal to the target (lungs, heart, and esophagus), excluding the spinal cord. Additionally, a 3mm planning organ at risk (PRV) spinal cord structure was generated to aid in decreasing spinal cord dose and to account for patient setup uncertainty.

**Table 1.** Ethos standard fractionation lung planning template. All optimization and planning organ at risk structures were generated in the Ethos workspace using the derivations summarized below.

| Priority | Structure | Structure Derivation | Planning Goal | Acceptable Variation |
| --- | --- | --- | --- | --- |
| 1 | PTVp | | V100% ≥ 98.5% | V100% ≥ 90% |
| | PTVn | | V100% ≥ 98.5% | V100% ≥ 90% |
| | PTVp | | V95% ≥ 100% | V95% ≥ 95% |
| | PTVn | | V95% ≥ 100% | V95% ≥ 95% |
| | Spinal Cord | | D0.03cc ≤ 30Gy | D0.03cc ≤ 50Gy |
| | PTVp | | D0.03cc ≤ 110% | D0.03cc ≤ 115% |
| | PTVn | | D0.03cc ≤ 110% | D0.03cc ≤ 115% |
| | _AvdLungs | Lungs – (PTV + 0.3cm) | V20Gy ≤ 25% | V20Gy ≤ 35% |
| 2 | SpinalCord_PRV03 | Spinal Cord + 0.3cm | D0.03cc ≤ 33Gy | D0.03cc ≤ 50Gy |
| | _AvdLungs | Lungs – (PTV + 0.3cm) | Dmean ≤ 15Gy | Dmean ≤ 20Gy |
| | _AvdHeart | Heart - (PTV + 0.3cm) | V30Gy ≤ 11% | V30Gy ≤ 50% |
| | _AvdHeart | Heart - (PTV + 0.3cm) | V45Gy ≤ 5% | V45Gy ≤ 40% |
| | _AvdEsophagus | Esophagus - (PTV + 0.3cm) | D0.03cc ≤ 57Gy | D0.03cc ≤ 63Gy |
| | _AvdEsophagus | Esophagus - (PTV + 0.3cm) | Dmean ≤ 28Gy | Dmean ≤ 34Gy |
| | Lungs-GTV | | V20Gy ≤ 28% | V20Gy ≤ 37% |
| | Lungs-GTV | | Dmean ≤ 20Gy | Dmean ≤ 22Gy |
| | Heart | | V30Gy ≤ 12% | V30Gy ≤ 55% |
| | Heart | | V45Gy ≤ 6% | V45Gy ≤ 40% |
| | Esophagus | | D0.03cc ≤ 63Gy | D0.03cc ≤ 66Gy |
| | Esophagus | | Dmean ≤ 34Gy | Dmean ≤ 40Gy |
| | _AvdLungs | Lungs – (PTV + 0.3cm) | V5Gy ≤ 55% | V5Gy ≤ 60% |
| | Lungs-GTV | | V5Gy ≤ 60% | V5Gy ≤ 65% |
| | Ipsilateral Brachial Plexus | | D0.03cc ≤ 66Gy | D0.03cc ≤ 66Gy |

The resulting template, generated on the tuning cohort, prioritized PTV coverage the most, followed by spinal cord avoidance, then PTV hotspot reduction. Lungs-GTV (referred to as lungs) V20Gy was deemed the most important OAR metric aside from the spinal cord D0.03cc (referred to as Dmax); its avoidance structure was therefore placed at the bottom of priority 1. All avoidance structures were assigned higher priorities than their respective OARs; this forced the template to prioritize target coverage over healthy tissue sparing, as the optimizer only avoided the entire OAR if it was at least 3mm from the target or all higher priority objectives have been met. The brachial plexus and esophagus were given the lowest and second lowest OAR priority, respectively. This template in XML format is included in Supplemental Materials.

### Plan geometry

The selected Ethos plans from the fifteen-patient tuning cohort include three equidistant 9-field plans (20%), four equidistant 12-field plans (27%), two ipsilateral 7-field plans (13%), one 2-arc VMAT plan (7%), and five 3-arc VMAT plans (33%). The plans selected from the validation cohort include nine equidistant 9-field plans (26%), thirteen equidistant 12-field plans (37%), eleven ipsilateral 7-field plans (31%), one 3-arc VMAT plan (3%), and one VMAT plan with 2 partial arcs (3%). Figure 1 shows Ethos and Eclipse dose distributions for validation cohort plans with the highest PTVp V100% coverage from each Ethos IMRT field geometry. Ethos 9- and 12-equidistant field IMRT plans avoid the spinal cord more than the Eclipse full arc plans, and the Ethos 7-lateral field IMRT plan avoids the heart more than Eclipse partial arc VMAT plan.

**Fig. 1.**
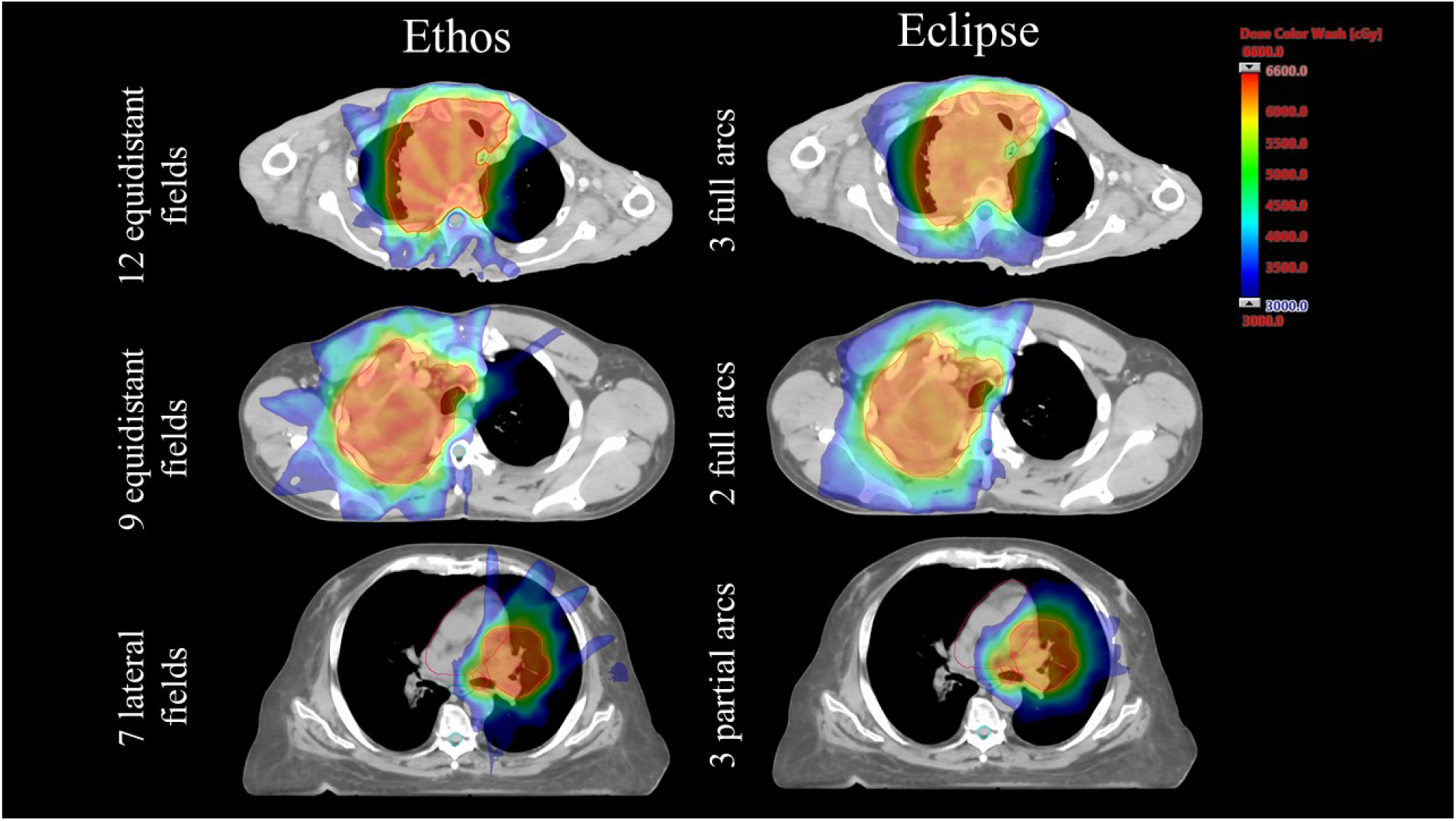
Ethos and Eclipse axial dose distributions for patients with the highest primary planning target volume (PTVp) V100% coverage for the three most prevalent Ethos field geometries (9 and 12 equidistant field intensity modulated radiation therapy (IMRT), 7 ipsilateral field IMRT). Identical slices are shown for both treatment planning systems (TPS). The PTV is contoured in red, the spinal cord in cyan, the heart in pink, and the esophagus in turquoise. The dose wash ranges from 30 Gy to 66 Gy, corresponding to dose levels between 50% and 110% of the prescription.

### Dosimetric comparison

The template automatically generated plans with clinically acceptable target and OAR metrics for 80% of tuning and validation patients per the ARTIA-Lung clinical trial. Figure 2 shows scatterplots comparing Ethos and Eclipse plan metrics for all ARTIA-Lung target and OAR objectives. To aid in visualization of metric distributions and outliers, box and whisker plots are shown for all ARTIA-Lung objectives in Supplemental Materials Figure A1. All PTVp, PTVn, spinal cord, heart, esophagus, and brachial plexus plan metrics were per protocol or variation acceptable without any further plan optimization. 20% (3/15) of the tuning cohort lungs V5Gy metrics were variation unacceptable, and 20% (7/35) of the validation cohort lungs V5Gy metrics were variation unacceptable. Of the 7 validation patients with unacceptable V5Gy values, one plan also contained an unacceptable lungs V20Gy metric.

**Fig. 2.**
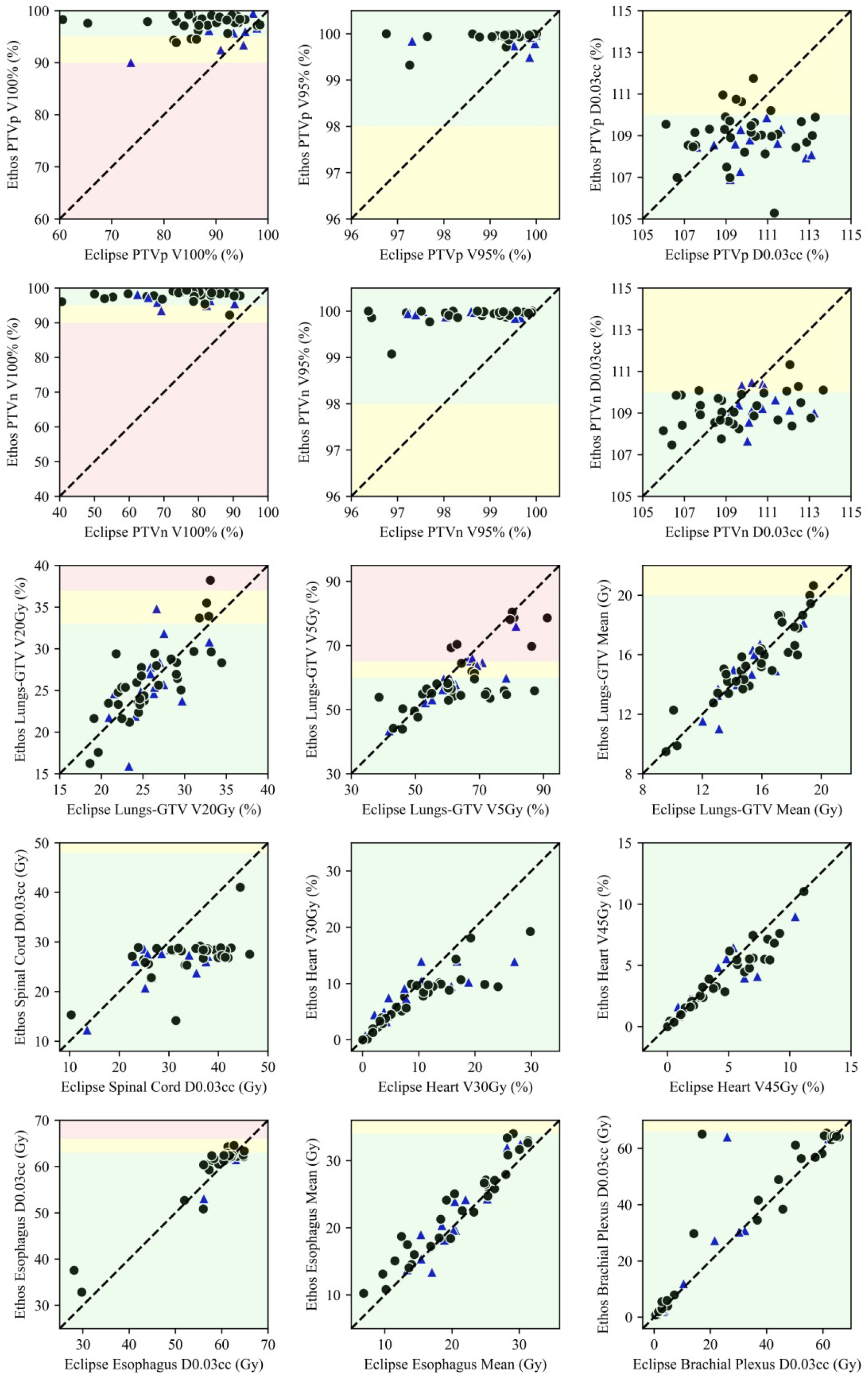
Scatter plots of all ARTIA-Lung clinical trial target and normal tissue metrics. Dashed lines indicate treatment planning system equality, data point shape specifies cohort (blue triangle = tuning, black circle = validation), and shaded regions illustrate constraint acceptability per the ARTIA-Lung clinical trial (green = per protocol, yellow = variation acceptable, red = variation unacceptable).

Table 2 shows mean and standard deviation values from the tuning and validation cohorts for all ARTIA-Lung trial metrics and all relevant QUANTEC metrics. To illustrate variation in Eclipse plan metrics as a function of recalculation alone, the originally planned AAA values and the recalculated AXB values are presented. Wilcoxon testing reveals significant differences between the Eclipse AXB and Ethos validation cohorts for most metrics. Ethos plans have significantly greater PTVp and PTVn V95% and V100% coverage (p<0.001 for all), and a lower Dmax for the PTVp (p=0.023). The average Ethos validation cohort spinal cord Dmax is about 7Gy less than that of Eclipse (p<0.001). While negligible difference is observed for the lung V20Gy and Dmean, Ethos V5Gy values are about 5% less than Eclipse, on average (p=0.003). Heart mean metrics are similar for both TPSs, but the heart V25Gy, V30Gy, and V45Gy are lower for Ethos (p=0.021, p<0.001, and p<0.001, respectively). Eclipse is higher than Eclipse for all esophagus (mean, V35Gy, V50Gy, Dmax) and brachial plexus (Dmax) metrics (p<0.001, p<0.001, p=0.002, p=0.001, and p=0.008, respectively).

**Table 2.** Summary of values for all XXX-XXX clinical trial dose volume histogram (DVH) metrics as well as pertinent Quantitative Analysis of Normal Tissue Effects (QUANTEC) DVH metrics not already listed. P values were obtained by performing a Wilcoxon signed rank test using the Eclipse and Ethos Acuros XB validation cohorts. P values < 0.05 are bolded and deemed significant.

| Structure | Dosimetric Parameter | Eclipse AAA |  | Eclipse AXB<br>(Recalculated) |  | Ethos AXB |  | P Value |
| --- | --- | --- | --- | --- | --- | --- | --- | --- |
|  |  | Tuning | Validation | Tuning | Validation | Tuning | Validation |  |
| PTVp | V100% (%) | 95.0 ± 3.4 | 95.6 ± 2.1 | 91.6 ± 6.1 | **87.2 ± 8.3** | 96.1 ± 2.7 | **97.5 ± 1.6** | **< 0.001** |
|  | V95% (%) | 99.9 ± 0.3 | 99.6 ± 0.6 | 98.9 ± 2.5 | **99.4 ± 0.9** | 99.9 ± 0.2 | **99.9 ± 0.1** | **< 0.001** |
|  | D0.03cc (%) | 108.5 ± 1.7 | 109.0 ± 1.5 | 110.4 ± 1.6 | **109.9 ± 1.9** | 108.5 ± 0.8 | **109.0 ± 1.2** | **0.023** |
| PTVn | V100% (%) | 93.2 ± 3.5 | 92.7 ± 2.9 | 78.6 ± 9.3 | **76.1 ± 12.2** | 97.5 ± 1.9 | **97.9 ± 1.5** | **< 0.001** |
|  | V95% (%) | 99.5 ± 0.6 | 99.5 ± 0.7 | 98.4 ± 1.2 | **98.6 ± 1.0** | 99.9 ± 0.1 | **99.9 ± 0.2** | **< 0.001** |
|  | D0.03cc (%) | 109.3 ± 1.6 | 109.4 ± 1.5 | 110.6 ± 1.0 | 109.5 ± 2.1 | 109.5 ± 0.8 | 109.2 ± 0.8 | 0.095 |
| Spinal Cord | D0.03cc (Gy) | 29.9 ± 6.4 | 35.6 ± 7.6 | 29.7 ± 7.3 | **34.1 ± 7.5** | 25.3 ± 4.2 | **27.3 ± 4.2** | **< 0.001** |
| Lungs -GTV | V20Gy (%) | 26.4 ± 3.0 | 26.7 ± 4.4 | 26.0 ± 3.0 | 26.0 ± 4.2 | 26.0 ± 4.6 | 26.3 ± 4.3 | 0.534 |
|  | V5Gy (%) | 63.9 ± 10.0 | 66.0 ± 13.1 | 63.2 ± 9.9 | **64.5 ± 12.9** | 59.6 ± 7.4 | **59.3 ± 9.0** | **0.003** |
|  | Dmean (Gy) | 14.8 ± 1.8 | 15.9 ± 2.6 | 14.6 ± 1.8 | 15.5 ± 2.5 | 14.5 ± 1.9 | 15.6 ± 2.4 | 0.857 |
| Heart | Dmean (Gy) | 10.2 ± 4.6 | 11.4 ± 5.9 | 9.8 ± 4.4 | 10.9 ± 5.7 | 10.4 ± 4.2 | 10.7 ± 5.3 | 0.053 |
|  | V25Gy (%) | 12.7 ± 8.7 | 15.6 ± 11.1 | 12.0 ± 8.5 | **14.7 ± 10.5** | 11.8 ± 6.0 | **13.0 ± 8.0** | **0.021** |
|  | V30Gy (%) | 9.7 ± 7.6 | 11.6 ± 8.6 | 9.2 ± 7.4 | **10.8 ± 8.1** | 7.9 ± 4.2 | **7.9 ± 4.2** | **< 0.001** |
|  | V45Gy (%) | 4.7 ± 4.7 | 4.4 ± 3.3 | 4.4 ± 4.5 | **4.1 ± 3.0** | 3.6 ± 2.6 | **3.6 ± 2.5** | **< 0.001** |
| Esophagus | Dmean (Gy) | 19.8 ± 6.4 | 21.9 ± 6.9 | 19.2 ± 6.2 | **21.1 ± 6.7** | 20.0 ± 7.0 | **23.0 ± 6.7** | **< 0.001** |
|  | V35Gy (%) | 27.4 ± 12.3 | 28.7 ± 14.9 | 26.5 ± 12.2 | **27.7 ± 14.9** | 27.8 ± 13.9 | **32.3 ± 15.2** | **< 0.001** |
|  | V50Gy (%) | 17.9 ± 10.3 | 18.1 ± 13.4 | 17.1 ± 10.1 | **17.3 ± 13.0** | 18.6 ± 11.6 | **20.0 ± 15.3** | **0.002** |
|  | D0.03cc (Gy) | 58.7 ± 11.9 | 59.0 ± 7.9 | 58.1 ± 11.9 | **58.6 ± 8.0** | 58.7 ± 12.4 | **59.8 ± 6.9** | **0.001** |
| Brachial Plexus | D0.03cc (Gy) | 13.4 ± 17.8 | 31.2 ± 26.3 | 13.3 ± 17.8 | **31.2 ± 26.7** | 16.3 ± 22.3 | **33.9 ± 27.5** | **0.008** |

Figure 3 shows validation cohort-averaged DVHs with standard deviation bounds for the PTVp, PTVn, spinal cord, lungs, heart, and esophagus. For comparison, tuning cohort averaged DVHs are shown in Supplemental Materials Figure A2. Ethos PTVp and PTVn have noticeably higher V100% values and steeper dose gradients than Eclipse, indicating greater target homogeneity.

**Fig. 3.**
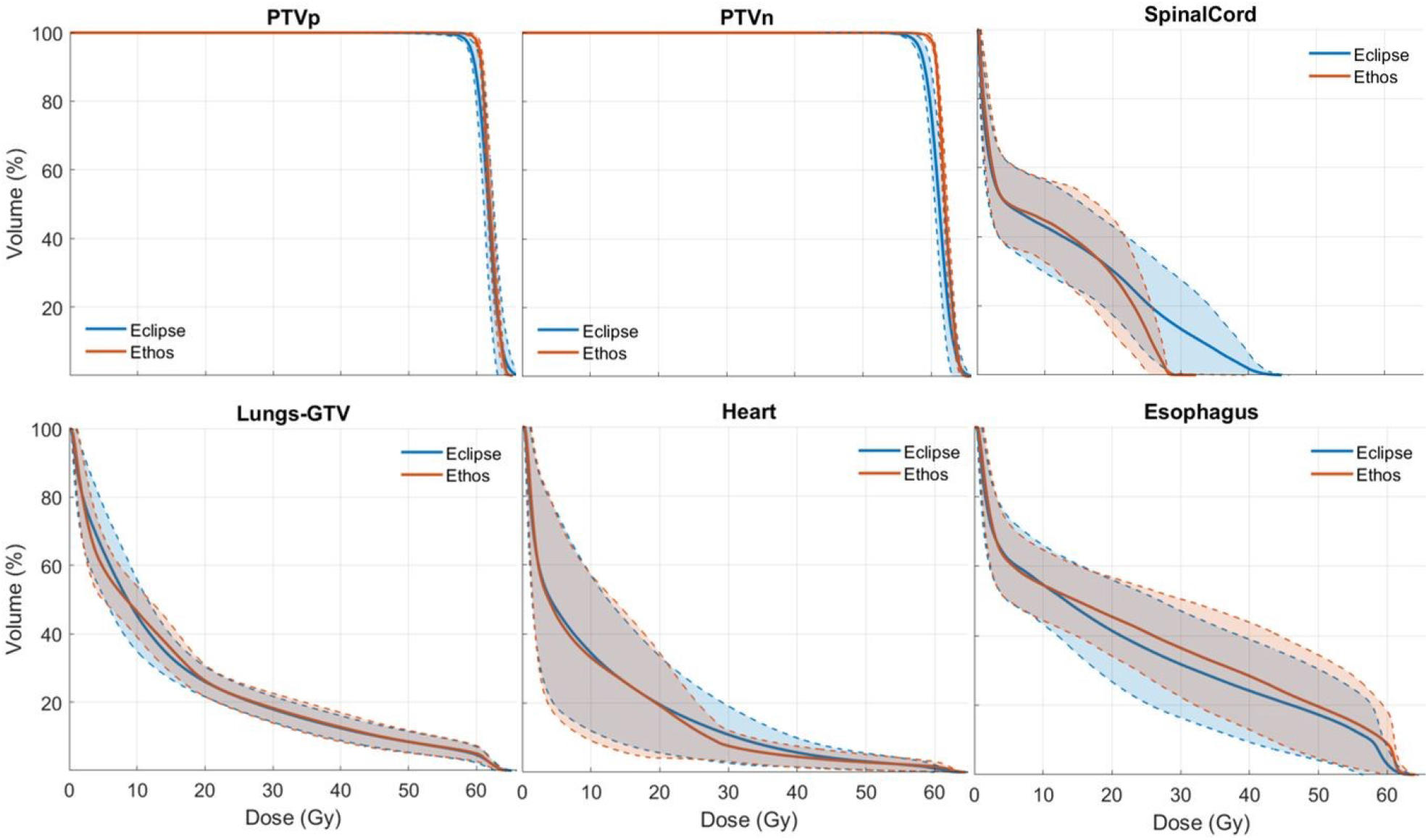
Cohort-averaged dose volume histograms for the primary and nodal planning target volume (PTV), spinal cord, lungs, heart, and esophagus for Ethos and Eclipse validation cohort plans. Solid lines represent the mean value and shaded regions illustrate all values within one standard deviation of the mean.

Ethos and Eclipse mean spinal cord dose diverges beyond 20Gy, with Ethos plans providing more sparing to the cord. Heart dose is only marginally reduced in Ethos plans from 0 - 20Gy, but the volume receiving dose above 20Gy is lower for Ethos plans. This is especially true for patients receiving above average heart dose. Ethos plans showed higher DVH values in the esophagus.

### Plan quality

As shown in Table 3, the mode score for physicians A and C is 5, while physician B has a mode score of 4. 78% of plans were considered of clinically acceptable quality to every reviewing physicians, without re-optimization, whereas 22% (11/50) of plans have at least one physician score of 3 due to hotspots in OARs or PTV undercoverage. No plans received a score of 1 or 2 from any physician. The average physician scores for the tuning and validation cohorts are 4.36 and 4.39, respectively, suggesting no decrease in quality when plans were generated without bias. While 22% of cases were deemed not clinically acceptable, the feedback from reviewers varied in some cases. For example, physician B gave one plan a score of 3 because coverage was too high, leading to a hotspots in the overlap of an OAR and PTV. However, physician A scored the same plan a 5, noting that the increased target coverage was preferred at the expense of higher brachial plexus dose. Physician C gave three plans scores of 3 based partially on suboptimal PTV coverage. However, in these three cases Ethos plans had greater PTV V100% and V95% metrics than Eclipse plans.

**Table 3.**
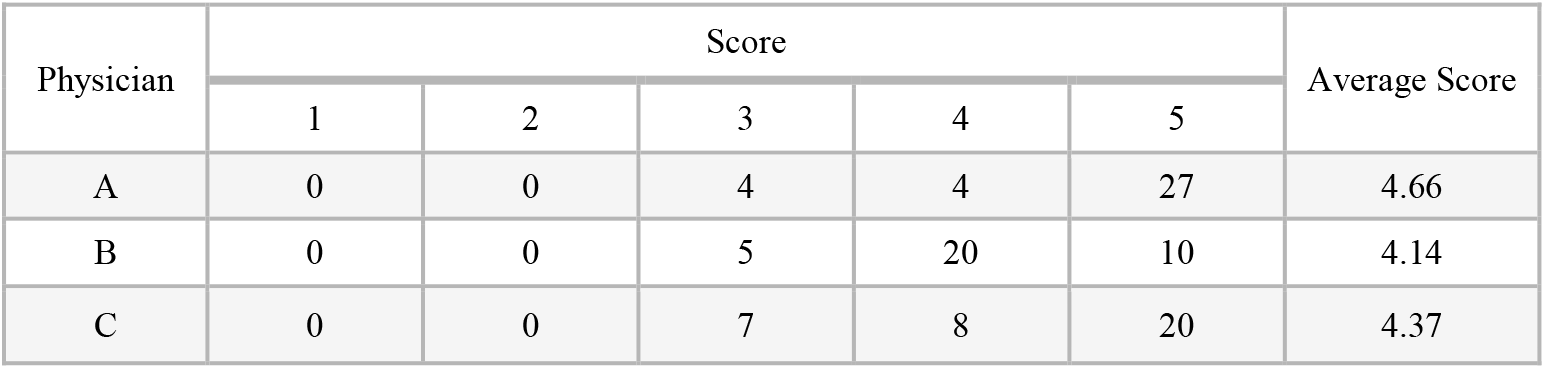
Qualitative plan review results by physician.

## Discussion

In this study we present, to our knowledge, the largest Ethos treatment planning study of a single treatment site, which evaluated automatically generated radiotherapy treatment plans for advanced lung cancer patients. Evaluation of the final planning template resulted in 80% of Ethos-generated plans meeting planning objectives without user input. These findings are supported by independent qualitative evaluation of individual plans, which showed that a large majority of plans (78%) lung would be approved by multiple physicians “as is” or with “minor edits that are not necessary” (i.e. stylistic in nature and likely not clinically impactful).

Interestingly, 40% of the selected tuning cohort plans were VMAT, but less than 6% of the selected validation cohort plans were VMAT. It is possible that the tuning cohort contained several outliers. However, the tuning cohort was randomly sampled from the fifty patient population such that an equal number of left and right disease sites were chosen (eight left, seven right). The high ratio of IMRT to VMAT plans in the validation cohort, along with the increase in time for VMAT dose calculation relative to IMRT, suggests that it may be clinically optimal to only perform IMRT calculations using this template, as adaptive treatment plans are calculated while patients are on the couch. These results are in good agreement with Calmels et al., who observed that, for most planning objectives, the Ethos TPS generates plans with similar or better target coverage and OAR sparing compared to manually generated Eclipse VMAT plans for anal, rectal, and prostate cancers.^19^ Thus, even though their study focused on pelvic sites, it indicates that one can expect an improvement in some, but not all, objective metrics when transitioning from manual Eclipse planning to automated Ethos planning.

It should be noted that further studies are needed to deconflate the effects of the Ethos treatment machine and optimizer on plan quality relative to Eclipse. More explicitly, a controlled experiment is required to determine what portion of Ethos plan difference is attributable to the double banked MLC, and what portion is attributable to its IOE. A potential avenue for future study entails directly comparing Ethos halcyon plans generated in Eclipse to those generated by the IOE.

While non-parametric testing is appropriate for analyzing difference in dose metric distributions that are not normal, non-parametric tests have no mechanism for capturing the magnitude of difference between two samples. As an example, the Wilcoxon signed rank test yields a significant value (p<0.001) when comparing Ethos and Eclipse PTVp V95% metrics because there is a high probability that a randomly sampled patient will have a V95% value that is lower in Ethos versus Eclipse. However, the means of the two V95% distributions are only 0.5% different. Thus, not all significant p values translate to clinically meaningful improvements when dealing with rank order tests. Because of this, DVH curve comparison (Figure 3) should be utilized to elucidate TPS dose difference continuously as a function of target/OAR volume when necessary.

For AAA, the PTV volume receiving prescription dose ranges from 86.7% - 96.7%. When all plans were recalculated in AXB, the range of PTV volumes receiving prescription dose decreases to 53.7% - 94.3%. While some of this discrepancy can be attributed to the fact that AAA overestimates dose to lung targets, ^21, 22^ it is likely that some decrease in target coverage occurs because re-optimization with an intermediate AXB calculation was not performed in Eclipse prior to recalculation with AXB. Despite this fact, statistical analysis shows that Ethos validation cohort PTVp and PTVn V100% metrics are greater than the originally calculated AAA metrics, not just the recalculated AXB metrics (p < 0.001 for PTVp and PTVn). Ethos plans are generated by optimizing the primary and nodal PTVs separately, allowing the targets to be uniquely adapted if the tumor and nodes respond differently during treatment. However, the majority of Eclipse plans only optimize using the total PTV. Separating the PTV into distinct primary and nodal structures gives the optimizer greater local dose control, providing another possible explanation for the increased Ethos target coverage relative to Eclipse.

The difference between Ethos and Eclipse lungs DVHs can be attributed to the characteristic difference in dose deposition between IMRT and VMAT; it is expected that the IMRT plans will result in a lower volume of lung receiving 0 – 10Gy due to dense volumes of dose at select gantry locations, also known as ‘streaking’. It is also expected that VMAT plans will result in a lower percent of lung volume receiving 10 – 20Gy due to a more homogeneous dose distribution. To the contrary, higher Ethos esophagus and brachial metrics are due to the architecture of the optimizer in the dose preview workspace. While all Eclipse structures can be given an equal priority and therefore have a similar effect on the cost function, Ethos optimizer priorities are necessarily different from each other. Therefore, esophagus objectives would need to be moved higher than spinal cord, lung, or heart objectives in the dose preview workspace to further improve Ethos esophagus metrics relative to Eclipse. However, the spinal cord, lungs, and heart were deemed more critical structures, and thus were given a higher priority. Even though the esophagus and brachial plexus metrics generated in the Ethos workspace were higher than Eclipse, all values were per protocol or variation acceptable, indicating the increase in OAR metrics is acceptable.

To investigate whether Ethos plans have increased esophagus and brachial plexus dose due to higher target coverage alone or due to the template itself, every Ethos plan was normalized so that the volume of PTV receiving prescription dose matched the clinically accepted Eclipse AAA plan. Following this normalization, Eclipse validation cohort esophagus dose is still lower than Ethos for all objective metrics. However, the Ethos PTVn D0.03cc and heart mean metrics become significantly lower than Eclipse (p=0.001 and p=0.029, respectively), and the Ethos brachial plexus metrics become similar to Eclipse (p=0.095) after normalization. Therefore, given equal prescription target coverage, this template generates Ethos plans with inferior esophagus metrics, similar brachial plexus Dmax, lung mean, and lung V20Gy metrics, and lower dose for every other metric of interest relative to Eclipse manually generated plans.

For Eclipse AXB plans, 40% (6/15) of the tuning cohort plans failed to meet the per protocol or variation acceptable lungs V5Gy objective and 43% (15/35) of validation cohort plans failed to meet these objectives. While the original clinical intent cannot be known retrospectively, nor were these plans generated according to the ARTIA-Lung clinical trial, it can be assumed that lung dose was limited to the extent possible during the initial planning process. Given 80% of Ethos plans were clinically acceptable, this suggests the Ethos template proposed in this work is well-tuned for limiting low levels of lung dose.

Several plan characteristics not evident by evaluating dose metrics alone were probed during physician review. Disease extent and locale were considered to ascertain whether each anatomy was favorable or challenging. Dose distributions were evaluated on a slice-by-slice basis, ensuring that the location of the dose inside the target or OAR was appropriate, not just the magnitude. The clinical impacts of high dose ‘streaking’ and hotspot location were also judged. Because physicians were not informed of the ARTIA-Lung clinical trial metrics prior to review, plan quality was judged using reviewer-specific target and OAR objectives which were subjective, and often more stringent than the trial objectives. Physicians were not satisfied with plans simply because all dose metrics were achieved, but were looking instead for techniques allowing plans to further improve. Contrary to template generated Ethos plans, 100% of Eclipse plans were assumed to be of clinically acceptable quality since they were each physician approved and treated clinically.

There is sizeable variation in score distributions among the reviewers, partially because no two physicians reviewed the same thirty-five plans, but mostly because every reviewer has subjective planning preferences. Generally speaking, physician A prefers target coverage over OAR sparing, physician B prefers minimizing maximum doses within OARs at the expense of target coverage, and physician C prefers that the target and OARs be somewhat equally prioritized.

Because the template in this work is biased towards the ARTIA-Lung clinical trial, potential users of the proposed template may find it beneficial to fine tune objective priorities and values to individual physician preferences, or instead to institutional standards and clinical trials if the template will be used more generally. To that end, the goal of this work is to elucidate planning techniques for generating high quality, automated lung plans in the Ethos workspace, not to propose that certain target and OAR constraints be implemented in a particular clinic. Additionally, the goal of the ARTIA-Lung trial is to reduce target margins for adaptive treatment. With smaller target margins there will be less overlap between OARs and targets, and thus less high dose to OARs.^30^ This template could therefore potentially generate superior plans for patients treated adaptively.

Future work includes the clinical implementation and prospective use of the resulting planning template. Furthermore, the methodology presented in this study will be translated to subsequent analyses focusing on extending our library of planning templates for other cancer sites.

The content and framework of this manuscript were constructed for consistency with the recently published radiation therapy treatment planning guidelines (RATING) for generating high quality planning studies.^31^ The agreed upon self-assessment score of two authors was 95% (196/207).

The resulting spreadsheet is shared in the Supplemental Materials.

## Conclusion

The Ethos planning template developed in this study was applied to fifty patients with stage IIIA-IIIC NSCLC. Automatically-generated plans improved target coverage and hotpots relative to the clinically delivered Eclipse plans with reduced maximum spinal cord dose as well as reduction in most heart and lung dose metrics of interest but with an increase in esophagus and brachial plexus dose. Automatically generated plans resulted in per protocol or variation acceptable dosimetry in 80% of cases, without intervention or replanning, and 78% of automatically generated plans were deemed clinically acceptable by multiple physicians. This template, or a variation thereof, enables an efficient, automated planning approach for patients requiring standard fractionation treatment for locally advanced lung cancer.

## Data Availability

All data produced in the present study are available upon reasonable request to the authors

